# Accelerated Metabolomic Aging and Its Association with Social Determinants of Health in Multiple Sclerosis

**DOI:** 10.1101/2025.01.29.25321260

**Authors:** Fatemeh Siavoshi, Rezvan Noroozi, Gina Chang, Vinicius A. Schoeps, Matthew D. Smith, Farren B.S. Briggs, Jennifer S. Graves, Emmanuelle Waubant, Ellen M. Mowry, Peter A. Calabresi, Pavan Bhargava, Kathryn C Fitzgerald

## Abstract

**Objectives:** Biological age may better capture differences in disease course among people with multiple sclerosis (PwMS) of identical chronological age. We investigated biological age acceleration through metabolomic age (mAge) in PwMS and its association with social determinants of health (SDoH) measured by area deprivation index (ADI).

**Methods:** mAge was calculated for three cohorts: 323 PwMS and 66 healthy controls (HCs); 101 HCs and 71 DMT-naïve PwMS; and 64 HCs and 67 pediatric-onset MS/clinically isolated syndrome patients, using an aging clock derived from 11,977 healthy adults. mAge acceleration, the difference between mAge and chronological age, was compared between groups using generalized linear and mixed-effects models, and its association with ADI was assessed via linear regression.

**Results:** Cross-sectionally, PwMS had higher age acceleration than HCs: 9.77 years in adult PwMS (95% CI:6.57–12.97, p=5.3e-09), 4.90 years in adult DMT-naïve PwMS (95% CI:0.85–9.01, p=0.02), and 6.98 years (95% CI:1.58–12.39, p=0.01) in pediatric-onset PwMS. Longitudinally, PwMS aged 1.19 mAge years per chronological year (95% CI:0.18, 2.20; p=0.02), faster than HCs. In PwMS, a 10-percentile increase in ADI was associated with a 0.63-year (95% CI:0.10-1.18; p=0.02) increase in age acceleration.

**Discussion:** We demonstrated accelerated mAge in adult and pediatric-onset PwMS and its association with social disadvantage.

## Introduction

Chronological age (cAge), defined by an individual’s birth date, plays an important role in the clinical course of multiple sclerosis (MS), influencing rates of disease progression and therapeutic response^1^. However, as MS course is heterogeneous across individuals, varying rates of MS disability progression are observed among people with MS (PwMS) of the same chronological age^2^. These variations are likely influenced by a complex interaction of factors that may include an intersection of genetic and environmental contributors, resulting in unique progression patterns and responses to treatment for each individual^3^. Biological age, a measure of physiological aging and cellular health, may better capture these underlying biological differences than cAge, offering a more accurate reflection of an individual’s health status.

Social and environmental conditions are critical in shaping these biological patterns. Social determinants of health (SDoH), which include non-medical factors such as where people are born, live, and grow up, as well as the quality of their social relationships and networks, influence both physical and mental health outcomes^4,5^. A growing body of research suggests that SDoH and related factors are strong contributors to disease outcomes and also the quality of life in individuals with MS^6,7^. In the general population, the early onset and heightened severity of age-related diseases among individuals with greater exposure to unfavorable SDoH suggest that such lived experiences may contribute to an accelerated pace of biological aging in these individuals^8,9^. Biological aging could thus be a pathway through which SDoH impacts MS disease progression, making it essential to examine whether a similar relationship exists in MS.

This study compares biological age, estimated through metabolomic profiles, in PwMS and healthy controls, and explores its association with SDoH in PwMS. While biological age is commonly assessed using various biomarkers, including telomere length, DNA methylation, or clinical laboratory values, quantifying biological age using metabolomics may provide additional insight^10–14^; metabolomic profiles reflect downstream products of complex biological processes influenced by the genome, transcriptome, proteome, and notably, a broad array of environmental exposures relevant to SDoH, offer a closer approximation of the aging phenotype^15^. Thus, metabolomic age (mAge) clock allows us to capture a broad spectrum of physiological changes, providing insights into cellular health and the cumulative impact of external and internal factors on aging in MS.

## Methods

### Study participants

Data for this study were pooled from participants in ongoing or completed clinical research studies at three sites, where blood samples were acquired for metabolomic analyses: the Johns Hopkins MS Precision Medicine Center of Excellence (JHU), the University of Miami Miller School of Medicine (UMMSM), and the pediatric MS Center at the University of California, San Francisco (UCSF). The JHU cohort included people with MS at various disease stages receiving disease-modifying therapies (DMT) of varying efficacy or no therapy and healthy controls (HCs). DMTs received at the time of blood draw were classified by efficacy as high-efficacy (natalizumab, ocrelizumab, rituximab, daclizumab), moderate-efficacy (fingolimod, dimethyl fumarate), or mild-efficacy (glatiramer acetate, interferon-β, teriflunomide, mycophenolate mofetil). The UMMSM cohort comprised MS patients who were DMT–naïve, within five years of symptom onset, and diagnosed under the 2017 McDonald criteria within two years. The UCSF cohort consisted of HCs and individuals with pediatric-onset MS (POMS) or clinically isolated syndrome (CIS) at risk of developing MS; all met the 2017 McDonald criteria, had clinical onset before age 18, and were within four years of symptom onset.

### Measure of social determinants of health

We used the area deprivation index (ADI) as the primary SDoH indicator at the census block group level. The ADI is a well-known composite index strongly linked to health outcomes, including biological aging in the general population and our previous work in PwMS. The ADI incorporates 17 different measures of socioeconomic status (SES) [e.g., income disparity, access to motor vehicles, occupational composition, single-parent household rate, divorce rate, English language proficiency, among others]^6,16,17^. For this analysis, we used the national ADI, which ranges from 1 (least disadvantaged) to 100 (most disadvantaged). Analyses linking the ADI and mAge include only the JHU cohort, as participant-level geocodes (used to derived census block groups) available at the time of blood sampling were only available for these individuals.

### Metabolomic measurements

All blood samples at each center were processed within 3 hours and stored at -80°C until metabolomics analysis, following a standardized protocol. For each of the cohorts, global untargeted metabolomic profiling was conducted at Metabolon Inc. (Durham, NC)^18^. Samples were first thawed and further prepared through a derivatization process. The derivatized samples were analyzed using either gas chromatography coupled with mass spectrometry or liquid chromatography paired with tandem mass spectrometry. The resulting mass spectra were compared to a reference library of known standards to identify individual metabolites accurately. The relative abundance of each metabolite was determined by the area under the curve from the generated mass spectra. Metabolomic profiling were conducted in five distinct batches for JHU, in two batches for UCSF, and in a single batch for the UMMSM cohort.

## Statistical analysis

### Preprocessing and quality control

Consistent quality control and harmonization steps were applied to ensure high data quality across cohorts. Initially, within each batch, metabolites with over 20% missing values across samples were excluded from further analysis. For the remaining metabolites, missing values were imputed using k-nearest neighbors (k=10) and log-transformed, yielding 332 and 646 metabolites consistently measured across all batches in JHU and UCSF, respectively, and 959 metabolites in UMMSM. Within-site data were harmonized across batches using ComBat, which is an empirical Bayes-based method originally developed for gene expression and previously applied by our team in the context of metabolomics^19^. Data were winsorized at the 99^th^ percentile. Finally, JHU and UCSF data were normalized using the Z-score normalization method to ensure consistency with prior metabolomics analyses incorporating multiple batches, and UMMSM data were scaled to a median of one, consistent with the normalization approach used in developing the mAge clock.

### Metabolomic age calculation

MAge was calculated using a previously published mAge prediction model based on healthy individuals aged 18-75 years from the INTERVAL study based in Cambridge, UK (n=11,977; 50.2% male), in which untargeted metabolomic profiling was similarly performed by Metabolon Inc^20^. In brief, the mAge model was developed using 826 metabolites (678 endogenous and 148 xenobiotics) using Ridge regression. The investigators developed two models: Model A (a more parsimonious model), including 826 metabolites (678 endogenous and 148 xenobiotics), and Model B, including 678 endogenous metabolites. To calculate the mAge for each cohort, we applied the intercept, sex coefficient, and the set of overlapping metabolites—332 in the JHU, 563 in the UCSF, and 731 in the UMMSM dataset—with Model A of the mAge clock. As our set of metabolites did not overlap completely with the set of metabolites in the original model, we excluded individuals with predicted implausible mAges (e.g., those <0 or >100; n = 0 for the JHU cohort and n = 0 for the UCSF cohort, n = 7 for the UMMSM cohort).

### Descriptive and analytical models

Descriptive analyses summarized categorical variables with frequencies and continuous variables with means, medians, and interquartile ranges as appropriate. To evaluate metabolomic aging in PwMS compared to HCs, we performed both cross-sectional and longitudinal analyses. Age acceleration, defined as the difference between mAge and chronological age for each individual, was the primary effect of interest.

For between-group comparisons in the UCSF and UMMSM cohort, which included one sample per individual, we applied generalized linear models. In the JHU dataset, which included multiple samples for 195 (50.13 %) individuals, we accounted for within-person correlation using a linear mixed-effects (LME) model. In exploratory analyses, for the subset of participants with multiple samples with follow-up exceeding one year (n=173), we fit an LME model assessing if PwMS exhibited a faster rate of metabolomic aging compared to HCs, including group (PwMS vs. HCs), time, and cross-product term; the group-time cross-product reflects a measure of whether metabolomic aging was increased in PwMS relative to HCs. The association between mAge and ADI in PwMS was assessed cross-sectionally using linear regression within the JHU cohort. We also explored whether the association between ADI and mAge is modified in different racial groups. All analyses were conducted using R version 4.4.1, with a significance threshold of P < 0.05.

## Results

### Demographics of the participants

Following the removal of mAge outliers, 389 participants with 683 samples from the JHU cohort, 131 from the UCSF cohort, and 172 from the UMMSM cohort were included in the analysis. In the JHU cohort, the median age of participants was 45.0 years (interquartile range [IQR]: 35.1– 53.8 years), with 72.6% female participants (n = 495 samples) and 82.0% identifying as White (n = 559 samples). Among samples from PwMS participants, 152 (28.1%) were from individuals with progressive MS. In the UCSF cohort, the median age was 14.7 (IQR=12.35- 16.32) years, with 47.33% female participants (n=62), and 85.50% identifying as White (n= 112). In the UMMSM cohort, the median age was 39 (IQR=33- 48) years, with 74.56% female participants (n=126). All participants in this cohort identified as White, had relapsing–remitting MS, and were DMT-naïve. The detailed characteristics of the included participants for the cohorts are shown in Table 1.

**Table 1.** Characteristics of included study participants.

| Variable | JHU<sup>1</sup> |  | UMMSM<sup>2</sup> |  | UCSF<sup>3</sup> |  |
| --- | --- | --- | --- | --- | --- | --- |
|  | HC | MS | HC | MS | HC | MS |
| No. of samples | 142 | 541 | 101 | 71 | 64 | 67 |
| No. of participants | 66 | 323 | 101 | 71 | 64 | 67 |
| No. of samples per participant (mean) | 2.15 | 1.67 | 1.00 | 1.00 | 1.00 | 1.00 |
| Follow-up time, mean (SD) | 2.38 (2.04) | 4.76 (2.74) | - | - | - | - |
| Age, y, mean (SD) | 42.54<br>(12.83) | 45.35<br>(11.90) | 47.26<br>(16.36) | 49.61<br>(13.21) | 14.30<br>(2.52) | 14.20<br>(2.72) |
| Age range (min, max) | 22, 84 | 18, 72 | 19, 76 | 18, 62 | 8, 18 | 7, 18 |
| Female sex, n (%) | 106 (74.65) | 390 (72.09) | 74 (73.27) | 55 (77.46) | 30 (46.87) | 32 (47.76) |
| Race |  |  |  |  |  |  |
| Black or African American, n (%) | 21 (14.79) | 70 (12.94) | 0 (0.00) | 0 (0.00) | 3 (4.69) | 5 (7.46) |
| White, n (%) | 105 (73.94) | 455 (84.10) | 101 (100) | 71 (100) | 56 (87.50) | 56 (83.58) |
| Others, n (%) | 16 (11.27) | 16 (2.96) | 0 (0.00) | 0 (0.00) | 5 (7.81) | 6 (8.96) |
| Progressive MS, n (%) | - | 152 (28.10) | - | 0 (0.00) | - | - |
| DMT Status<sup>4</sup> |  |  |  |  |  |  |
| No DMT, n (%) | - | 102 (18.85) | - | 71 (100) | - | 12 (17.91) |
| Mild efficacy, n (%) | - | 191 (35.30) | - | 0 (0.00) | - | 5 (7.46) |
| Moderate efficacy, n (%) | - | 63 (11.65) | - | 0 (0.00) | - | 0 (0.00) |
| High efficacy, n (%) | - | 185 (34.20) | - | 0 (0.00) | - | 1 (1.49) |
| Missing, n (%) | - | - | - | - | - | 49 (73.14) |

1. JHU: Johns Hopkins MS Precision Medicine Center of Excellence
2. UMMSM: University of Miami Miller School of Medicine
3. UCSF: University of California, San Francisco
4. Disease-modifying therapy (DMT) status reflects the treatment participants were receiving at the time of the blood draw, classified as high-efficacy (natalizumab, ocrelizumab, rituximab, daclizumab), moderate-efficacy (fingolimod, dimethyl fumarate), mild-efficacy (glatiramer acetate, interferon beta, teriflunomide, mycophenolate mofetil), or not on DMT.

### Metabolomic age comparison

Significant differences in mAge acceleration between PwMS and HCs were observed; Cross-sectionally, adult PwMS exhibited a pronounced increase in age acceleration, with a mean difference of 9.77 years (95% CI: 6.57–12.97; p=5.3e-09) compared to HCs in the JHU cohort and 4.90 years in the UMMSM cohort (95% CI: 0.85–9.01; p=0.02). In the JHU cohort there was no overall effect of DMT class on mAge acceleration (p=0.33). Participants with POMS/CIS demonstrated an average increase in age acceleration of 6.98 years (95% CI: 1.58–12.39; p=0.01) compared to HCs (Figure 1).

**Figure 1.**
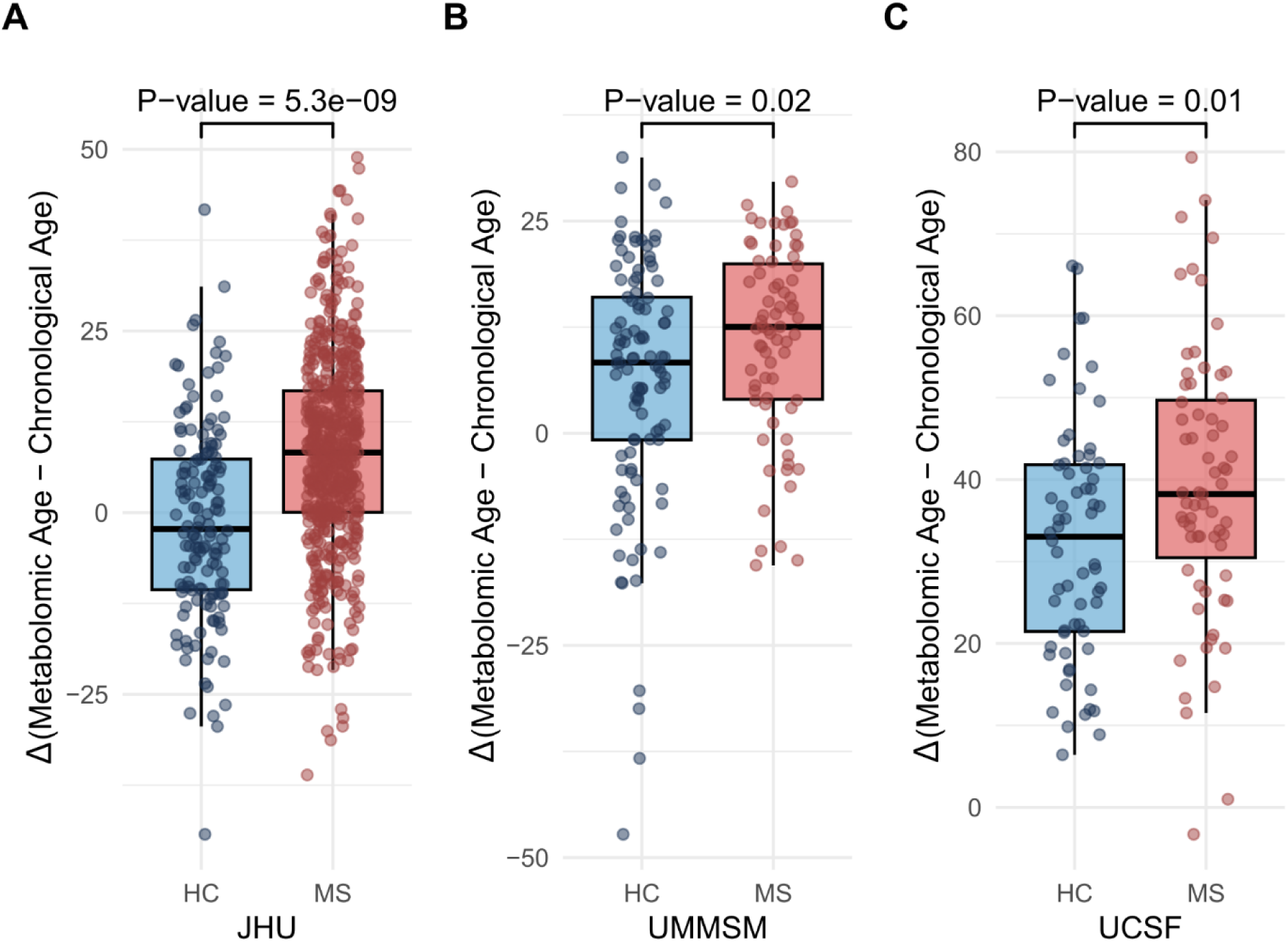
Metabolomic aging in multiple sclerosis. Comparison of metabolomic age acceleration (metabolomic age - chronological age) between individuals with Multiple Sclerosis (MS) and Healthy Controls (HC) across cohorts. **A.** Johns Hopkins MS Precision Medicine Center of Excellence (JHU) cohort **B.** University of Miami Miller School of Medicine (UMMSM) cohort **C.** University of California, San Francisco (UCSF) cohort

Longitudinal analysis in participants with follow-up data (mean follow-up: 4.57 years) in the JHU cohort revealed a faster metabolomic aging rate in PwMS than in HCs, with an average increase of 1.19 mAge years per chronological year (95% CI: 0.18, 2.20; p=0.02).

### Correlation of metabolomic age acceleration with ADI

ADI ranged 1 to 100 with a mean (SD) of 27.1 (20.7). Increasing social deprivation was associated with accelerated aging, with faster metabolomic aging. In PwMS, a 10-percentile increase in ADI was associated with a 0.63-year (95% CI: 0.10, 1.18; p=0.02) increase in age acceleration in PwMS after accounting for chronological age, gender, race, MS subtype, and MS therapy (Figure 2). We did not note effect modification by race (p=0.88).

**Figure 2.**
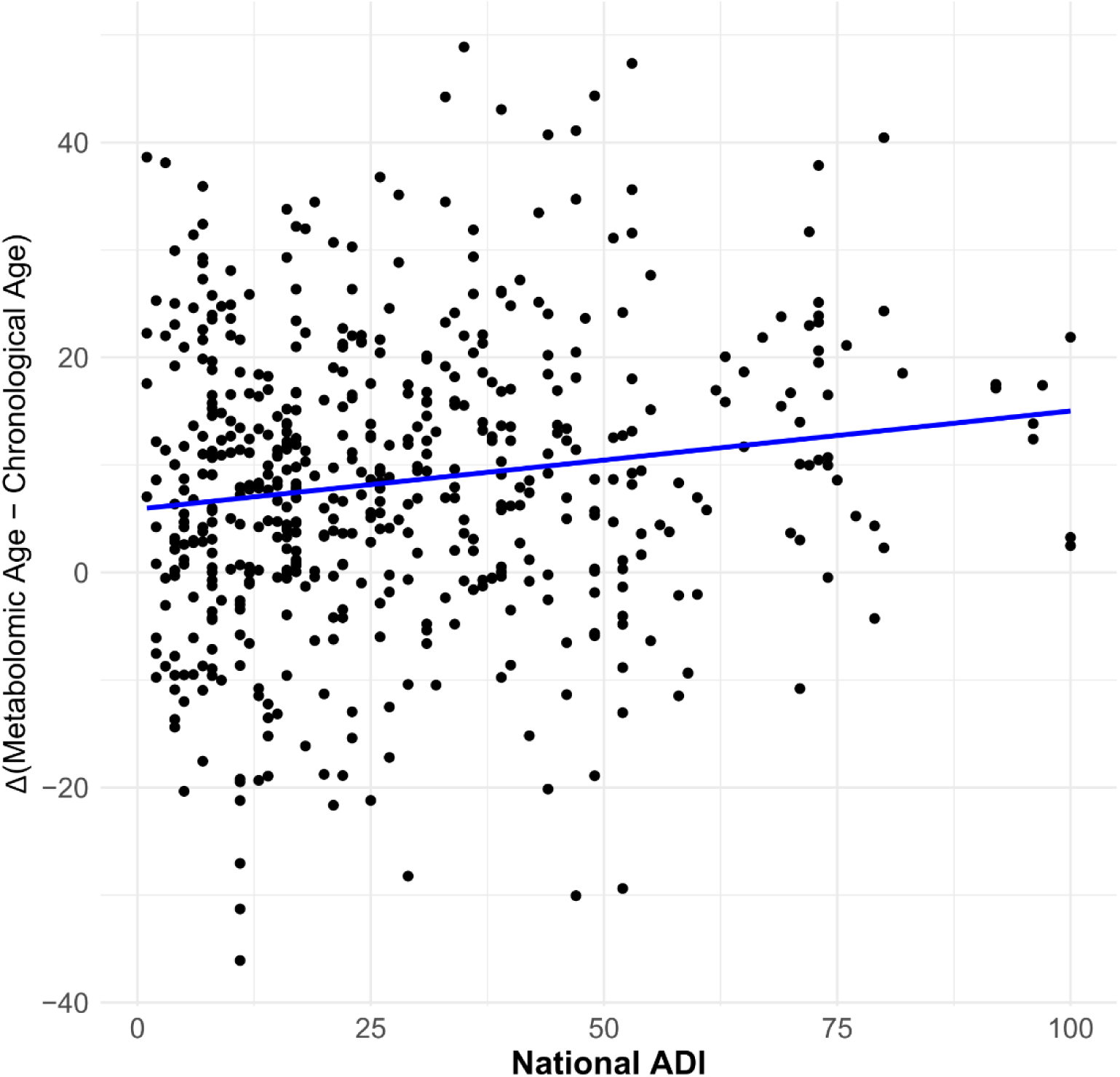
Relationship between metabolomic aging and social determinants of health in multiple sclerosis. Association of social deprivation with metabolomic age acceleration in people with MS in the JHU cohort

## Discussion

Our study demonstrates that PwMS experience accelerated metabolomic aging compared to HCs, as shown by mAge analyses conducted in both cross-sectional and longitudinal assessments. These findings are particularly important, as evidence of age acceleration was evident even in pediatric-onset MS and in DMT-naïve patients early in their disease course. This observation underscores that accelerated aging effect in MS begins early in the disease course and is independent of advanced disease progression or prolonged DMT exposure, highlighting the importance of interventions targeting aging in MS management.

Our findings are consistent with a growing body of research indicating that PwMS exhibit signs of premature aging across various biological systems, including elevated markers of oxidative stress, telomere shortening, and neuroinflammation^12,21^. By leveraging a metabolomic clock rather than single-marker approaches such as telomere length or composite clinical indices, we captured a multidimensional portrait of these aging hallmarks, integrating aging-related molecules and environmental exposures, into a single measure^15^. The accelerated aging in MS likely arises from chronic inflammation, immune dysregulation, and cellular stress. These factors, inherent to MS pathology, may drive biological processes typically associated with aging, such as mitochondrial dysfunction and DNA damage^22,23^. This accelerated aging may contribute to the early onset of age-related comorbidities, including cardiovascular disease and cognitive decline observed in PwMS, vulnerabilities that appear to be further intensified by social deprivation^1,24–26^.

Our study found that greater social deprivation, was associated with even faster metabolomic aging among PwMS. This link may suggest that socioeconomic factors can amplify the biological effects of MS, potentially accelerating aging processes and worsening disease progression. The ADI encompasses multiple aspects of social deprivation, such as income, education, employment, and housing quality. Each of these dimensions may contribute to chronic stress or limit access to resources essential for health maintenance, and disrupted dietary patterns, creating an added burden that may expedite biological aging^27^. Lower SES and greater social deprivation are associated with worse disease progression and higher disability levels in MS, and our results provide biological insight into this relationship^6,7^.

Accelerated biological aging may be one possible pathway through which social deprivation influences health, creating a compounding effect that potentially intensifies MS severity and increases disability risk. The identification of biological age as a mediator is consistent with cardiovascular studies demonstrating social factors impact health outcomes through accelerated aging^9,28^. Confirmation that accelerated aging mediates the association between social deprivation and MS outcomes would underscore the need to incorporate social determinants as modifiable risk factors in MS management—improving immediate health outcomes and mitigating long-term disease progression, specifically for lower SES populations. Furthermore, mAge acceleration could serve as a measurable target for intervention strategies aimed at reducing health disparities.

This study has several limitations worth noting. The optimal approach for calculating mAge would involve using the full set of metabolites from the mAge model. However, due to the study’s retrospective design, we were unable to utilize the complete set of metabolites included in the mAge clock to calculate mAge in cohorts, leading to differences in the metabolites used for each. Consequently, a direct comparison of mAge values between the cohorts, which differ in demographic and clinical characteristics, was not feasible. Because no metabolomic aging clocks exist for pediatric populations and to maintain consistency across cohorts, we applied the adult-trained clock to the pediatric samples. As a result, absolute mAge values in the UCSF cohort are inflated relative to chronological age, and age-acceleration measures should be interpreted only as relative differences between groups rather than true mAges. Additionally, longitudinal data and data necessary for calculating the ADI score were unavailable for participants in the UMMSM and UCSF cohorts, limiting our ability to generalize ADI-associated findings in these cohorts. While we used the ADI as a proxy for SES, its modest effect size on age acceleration may reflect that ADI does not fully capture the broader spectrum of SDoH at the individual level, such as social support, healthcare access, and lifestyle factors, which could also impact metabolomic aging. Although no significant racial differences were observed in the relationship between mAge acceleration and ADI, the predominance of White participants in the JHU cohort may have limited power to detect race-specific effects, and the generalizability of these findings. Expanding future analyses to include additional social variables and more racially diverse cohorts would offer a more comprehensive understanding of the relationship between SDoH and metabolomic age.

Prospective studies in more racially diverse cohorts using the full mAge model and detailed socioeconomic data could enhance the precision and generalizability of these findings.

## Conclusion

In conclusion, our study demonstrated accelerated metabolomic aging and its association with social disadvantage in PwMS. While these findings offer valuable insights into how socioeconomic disadvantage may contribute to MS severity and progression, future studies with a prospective design that utilizes a more comprehensive set of metabolites, alongside a broader range of social determinants, will help further validate these findings and unravel specific mediators of accelerated aging.

## Acknowledgments

This work was supported by several grants (JF-2007-36755 and R21NS123141 to PB, R01NS117541 to EW, R01NS133005 to KCF, R01NS121928 to FBSB). The authors are grateful to all the patients and healthy individuals who participated in the study.

## Author Contributions

PB and KCF contributed to the conception and design of the study. FS, RN, VS, MS, GC, PB, KCF, EW, JSG, FBSB, EMM, and PAC participated in the data acquisition, analysis, and manuscript editing. FS, PB, and KCF contributed to the drafting of the manuscript. All authors read and approved the final manuscript.

## Ethical consideration

Human research approval was obtained from the Institutional Review Boards at Johns Hopkins University, University of Miami Miller School of Medicine, and University of California, San Francisco, with ethical approval.

## Consent to participate

Written informed consent was obtained from all participants for participation and for the use of their blood samples in metabolomics analyses.

## Consent for publication

Not applicable

## Data Availability

The study’s anonymized data will be available from the corresponding author upon reasonable request.

